# Psychometric Properties of WHO’s Schedules for Clinical Assessment in Neuropsychiatry: A Systematic Review

**DOI:** 10.1101/2025.09.13.25335691

**Authors:** Wubalem Fekadu, Awoke Mhiretu, Atalay Alem, Traolach Brugha, Mark van Ommeren, Somnath Chatterji, Charlotte Hanlon, Abebaw Fekadu

**Author notes:** Corresponding author, *P.O.BOX: 9086.

## Abstract

**Background:** WHO’s Schedules for Clinical Assessment in Neuropsychiatry (SCAN) is often used as the gold standard for psychiatric classification. We systematically reviewed studies on the psychometric properties of the SCAN to support its adaptation to the revised international classification systems.

**Methods:** We searched PubMed, PsycINFO, Embase, Global Health, and Global Index Medicus up to April 17, 2025, and contacted experts. The protocol was registered in PROSPERO (CRD42024522395).

**Results:** Titles and abstracts of 4,241 records were screened, with 296 full-text articles evaluated. Ninety-three articles were included in the final review: 46 assessing SCAN’s psychometric properties and 47 validating other measures using SCAN as a gold standard. The internal consistency of the SCAN and its predecessor, the Present State Examination (PSE), ranged from good to excellent. Both demonstrated acceptable intra-rater, inter-rater, and test-retest reliability, with reliability especially high for psychotic disorders. There was also evidence supporting concurrent, construct, semantic, and content validity, although there was an absence of evidence for predictive validity. We also found acceptable psychometric properties for the different syndrome-based sections of the SCAN.

**Conclusion:** Although recent, high-quality studies are scarce, the SCAN is a promising tool for diagnosing a variety of psychiatric issues, particularly psychotic disorders. It demonstrates established reliability and evidence of concurrent, construct, semantic, and content validity. However, there is a need to revise the current version of SCAN to align it with contemporary diagnostic systems. Additionally, further research is required, especially regarding the assessment of non-psychotic conditions.

## 1. Introduction

Structured mental disorders’ classification instruments remain fundamental tools for mental health research and evidence-based practice. While simple self-reported symptom scales are commonly used to determine the occurrence of mental health symptoms and track the course of illness, classification tools are critical for accurately identifying illness conditions, delineating co-morbidity, and assisting in planning services and better care (Sheehan et al., 1998).

The Schedules for Clinical Assessment in Neuropsychiatry (SCAN) is a semi-structured World Health Organization (WHO) assessment interview tool that systematises the clinical examination, the elicitation of psychopathology to match a standard definition in a glossary, and the classification of neuropsychiatric conditions (WHO, 1994; J. Wing, Nixon, Mann, & Leff, 1977; J. K. Wing et al., 1990). SCAN allows the identification of disorders according to standard international diagnostic systems, through accompanying algorithms that interpret the clinical descriptions or diagnostic criteria of these systems, and offers the possibility of dimensional categorization of psychopathology (WHO, 1994) .

The development of SCAN began in the late 1950s, with the Present State Examination, and has since been revised several times. These revisions have remained true to the fundamental commitment of the SCAN to ensuring accurate assessment, systematisation, and transcultural applicability (Maurer, Hillig, Freyberger, & Velthaus, 1991; Rijnders et al., 2000; J. Wing et al., 1977; J. K. Wing et al., 1990). It has been translated into more than 35 languages (WHO, 1994)

SCAN is a collection of instruments designed to assess, measure, and classify psychopathology. It consists of four main components: the Present State Examination, the Glossary of Differential Definitions, the Item Group Checklist, and the Clinical History Schedule. The Present State Examination is divided into two parts: Part One focuses on non-psychotic disorders, while Part Two addresses psychotic and cognitive disorders (WHO, 1994).

The SCAN offers flexibility in constructing a clinical database which is not confined to any specific nosology. This approach enables adaptability in the interview process, allowing for modifications in question order and wording based on the course of the interview. Interviewers have the freedom to explore certain lines of inquiry while bypassing others as needed. Importantly, the examiner determines the presence of symptoms rather than relying solely on the person’s report.

SCAN has been used in efforts to examine the manifestations and epidemiology of psychosis in the Global South (Hanlon et al., 2024; Morgan et al., 2023) and for trials of interventions for people with psychosis in Africa (Hanlon et al., 2016). It was also used in Indigenous Andean culture (Uscamayta Ayvar et al., 2022). SCAN is used every seven years in England in the Adult Psychiatric Morbidity Survey (APMS) as a second-stage (confirmatory) examination in persons in the community when psychosis is suspected (Qassem et al., 2015).

It is being utilized in a global study across 19 sites in 12 countries throughout Asia, Africa, the Americas, and Europe. The aim is to understand the molecular and systemic factors linking COVID-19 to both short-term and long-term neurological illnesses (de Erausquin et al., 2022). Although studies have evaluated the psychometric properties of SCAN (Rijnders et al., 2000; Schützwohl, Kallert, & Jurjanz, 2007a) or some of its sections (Krisanaprakornkit, Rangseekajee, Paholpak, & Khiewyoo, 2007), there are also questions about its design, the requirement for specialized training, and the perceived unfriendliness of the format (Sheehan et al., 1998). Moreover, with the availability of new diagnostic instruments since the development of the PSE (and the SCAN), for example, the Structured Clinical Interview for DSM (SCID), it is important to revisit its ongoing relevance in a way that informs the SCAN revision for ICD-11 and the broader advancement of psychiatric research enabled by technology.

We conducted a systematic review to establish the psychometric properties of the various revisions of the SCAN, including its predecessor, the Present State Examination (PSE). We also profiled the studies that used the SCAN as a gold standard in validating other diagnostic and screening measures.

## 2. Methods

We adhered to the updated Preferred Reporting Items for Systematic Reviews and Meta-Analyses (PRISMA) guideline to conduct and report this systematic review (Page et al., 2021) and the COnsensus-based Standards for the selection of health Measurement INstruments (COSMIN) criteria (Prinsen et al., 2018). These guidelines cover reporting, data extraction, methodological quality criteria, and risk of bias assessment (Gagnier, Lai, Mokkink, & Terwee, 2021). The protocol was registered on the PROSPERO International Register of Systematic Reviews (CRD42024522395).

### 2.1. Search strategy

We searched five databases: PubMed, Embase, PsycINFO, Global Health, and Global Index Medicus. The databases were searched from the inception of each database with no language restriction up to April 17, 2024. Forward and backward searches were conducted to identify additional relevant studies. We also ‘snowballed based on recommendations of people who had published using SCAN The search terms consisted of keywords, MeSH and Emtree terms, and vocabularies for SCAN and psychometric properties. The terms were combined with the Boolean term AND:

#### Terms

1. **Terms for SCAN:** SCAN OR SCAN 1·0 OR SCAN 2 OR SCAN 2·1 OR “Schedules for Clinical Assessment in Neuropsychiatry“ OR “Schedules for Clinical Assessment in Neuropsychiatry 1.0“ OR “Schedules for Clinical Assessment in Neuropsychiatry 2.0“ OR “Schedules for Clinical Assessment in Neuropsychiatry 2.1“ OR PSE OR “Present State Examination” OR “MINI SCAN”
2. **Terms for psychometric Properties:** Psychometric* OR Validity OR Reliability OR “Reproducibility of Results” (Supplementary file_1).

### 2.2. Eligibility criteria

We included studies that met the following criteria:

1. Validation studies that reported the psychometric properties of any of the versions of the SCAN;
2. Studies that used SCAN as a gold standard to validate other diagnostic and screening measures.

### 2.3. Data extraction

WF and AM screened the titles and abstracts of identified records. Two reviewers (WF and AM) then independently conducted full article reviews. Discrepancies were resolved with discussion. The excluded articles and reasons for exclusion were documented. The two reviewers divided the studies and extracted data, frequently discussing them when needed. The data extraction was started on May 6^th^ 2024. The extraction sheet included; author, publication year, country, the SCAN sections assessed, participant characteristics and psychometric properties.

### 2.4. Risk of Bias Assessment

The two reviewers assessed bias using the COSMIN checklist (L. B. Mokkink et al., 2018). The tool has 10 domains of psychometric properties: tool development/adaptation procedure, content validity, structural validity, internal consistency, cross-cultural validity, reliability, measurement error, criterion validity, hypotheses testing for construct validity, and responsiveness. Items under each domain are rated as very good, adequate, doubtful, and inadequate.

### 2.5. Data synthesis

We summarized the results of studies on SCAN’s psychometric properties using descriptive statistics. We narrated the findings for domains where a statistical summary was not applicable. We narrated the profile of the studies that used SCAN as a gold standard, including the type of instrument, setting, and country.

## 3. Results

### 3.1. Study selection

See Figure 1 for the PRISMA diagram. A total of 4,241 records were included. After title and abstract screening, 3,977 articles were removed because they did not study any of the SCAN versions. Two hundred ninety-six articles were considered for full-text review, including 32 articles from experts’ suggestions and hand-searched similar articles. Out of these, 203 were not eligible for this review because the studies mainly used SCAN for diagnosing neuropsychiatric problems for prevalence or intervention studies, rather than evaluating the properties of SCAN. In total, 93 articles were included in the review: 46 assessed the psychometric properties of SCAN, while the remaining 47 articles used SCAN as a gold standard.

**Figure 1:**
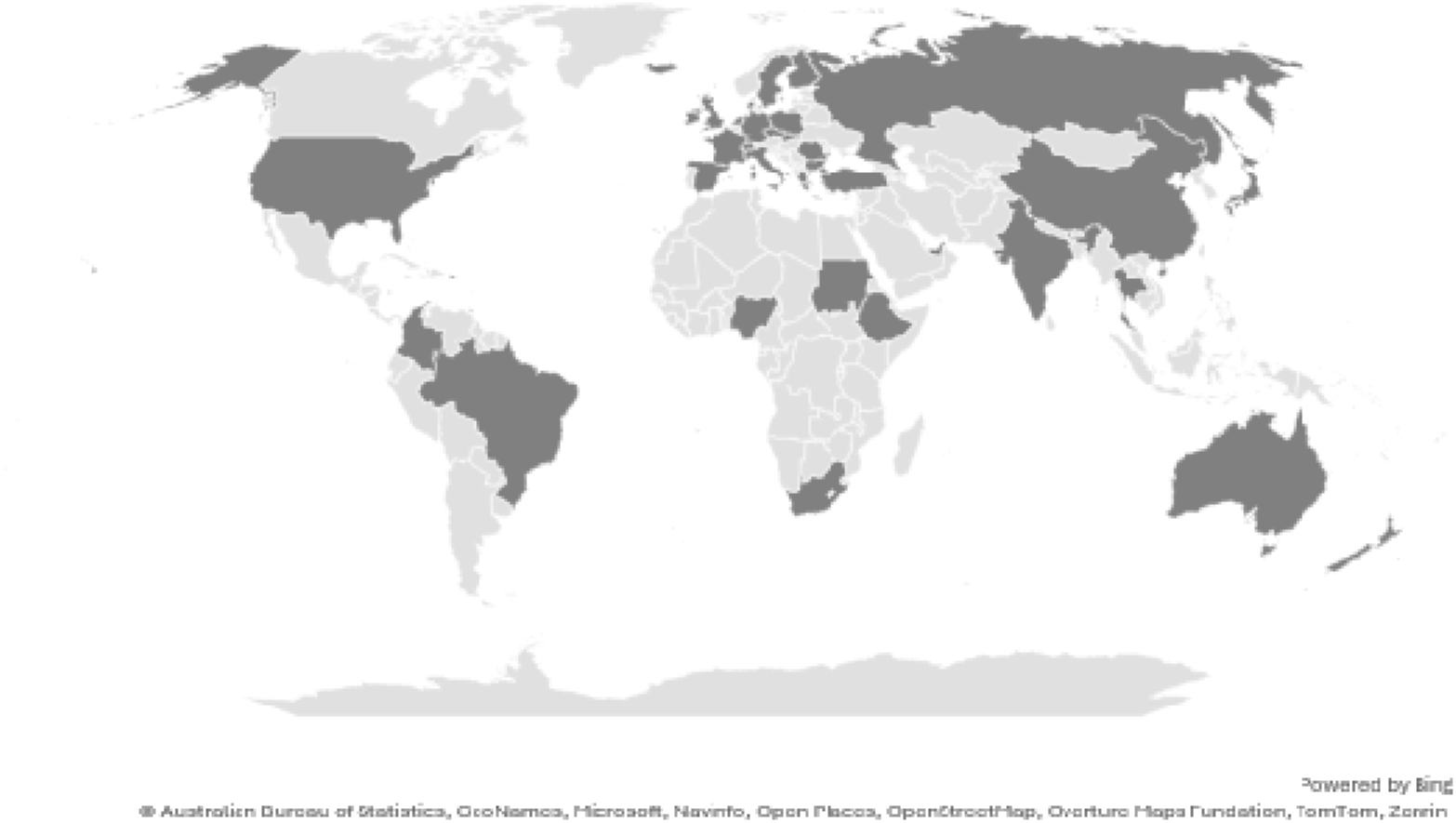
Countries where SCAN has been used

### 3.2. Study characteristics

Among the 46 articles assessing the performance of SCAN, 31 articles were from high-income countries. Two were multi-county studies that included at least one low- and/or middle-income country (Ustün et al., 1997; Vázquez-Barquero et al., 1994). Out of the remaining 13, nine studies were from Thailand, two were from Ethiopia, one from Turkey and one from Brazil. In these studies, SCAN and PSE were translated into more than 19 languages. The publication year ranged from 1977 to 2011 (Figure 2).

**Figure 2:**
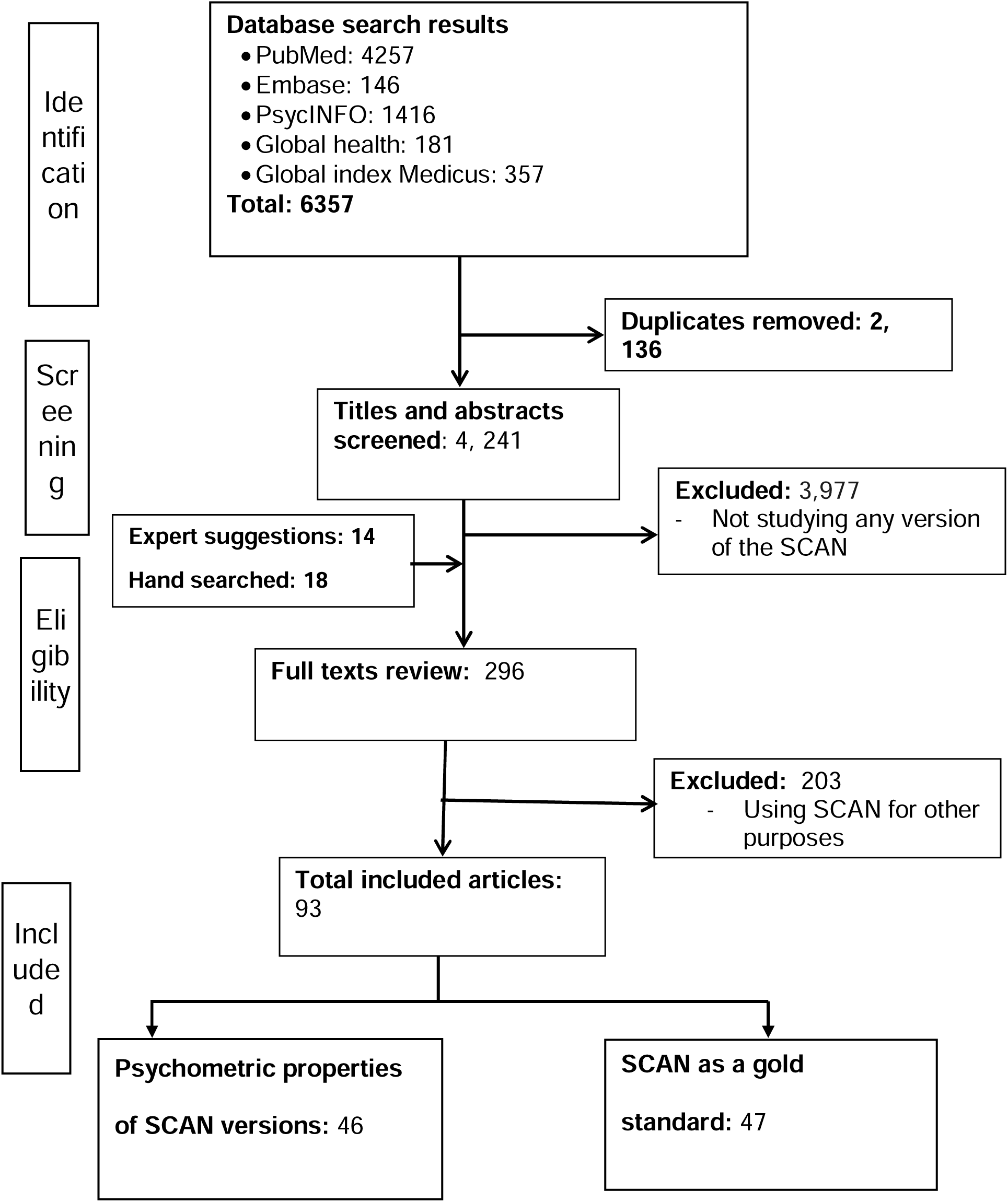
PRISMA flow diagram of the study selection process

The study sample sizes ranged from eight video-recorded ratings to 1244 general population participants. These studies were conducted among participants admitted to hospitals, outpatient departments, and general population settings. Some of the studies recruited participants from more than one place.

Twenty-eight studies reported on the full SCAN or PSE, while the remaining 18 studies reported on selected sections or items. Most of the studies used either trained psychiatrists, general practitioners or clinical psychologists, while some employed trainee psychiatrists, psychiatric nurses and lay interviewers (Table 1)

**Table 1:**
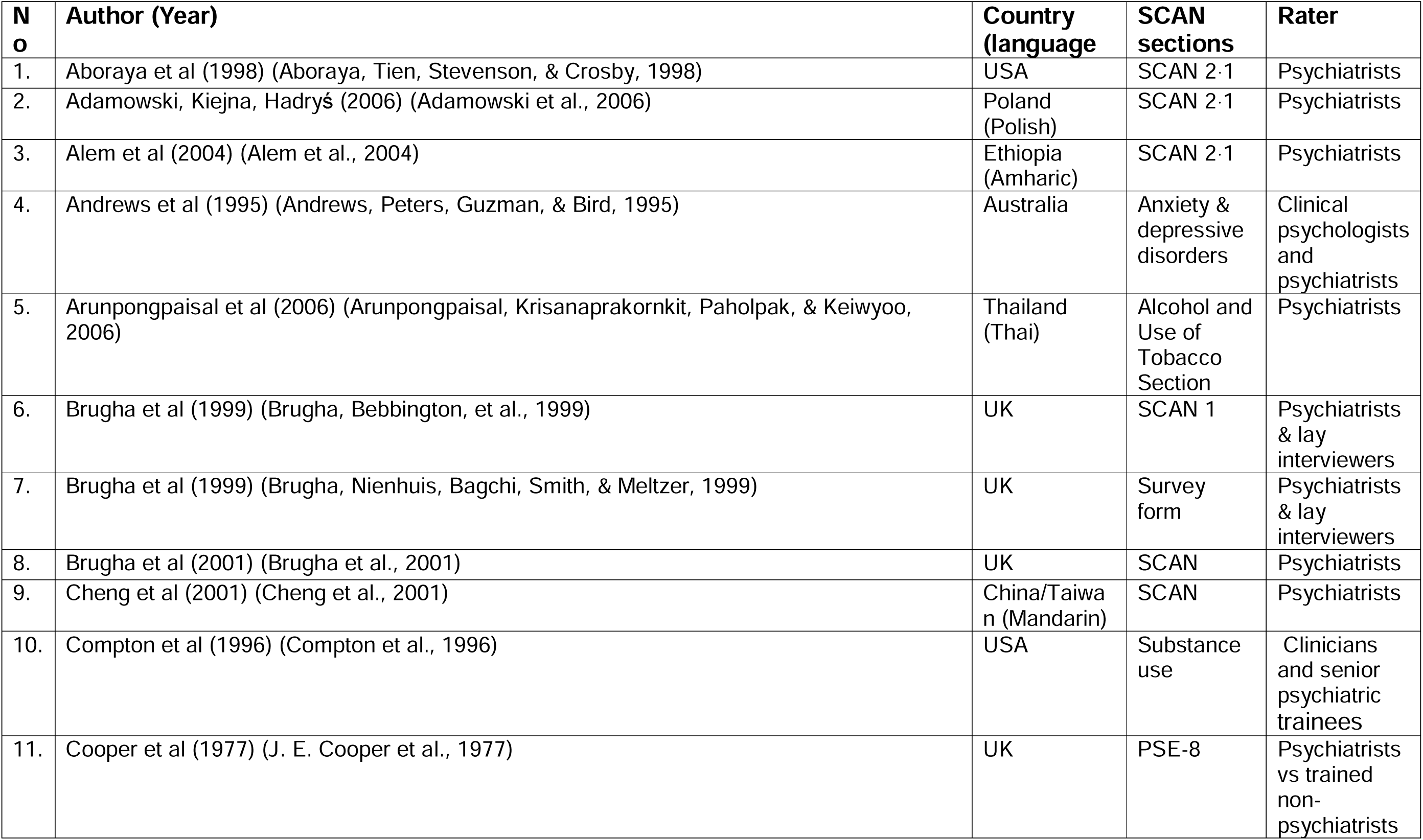
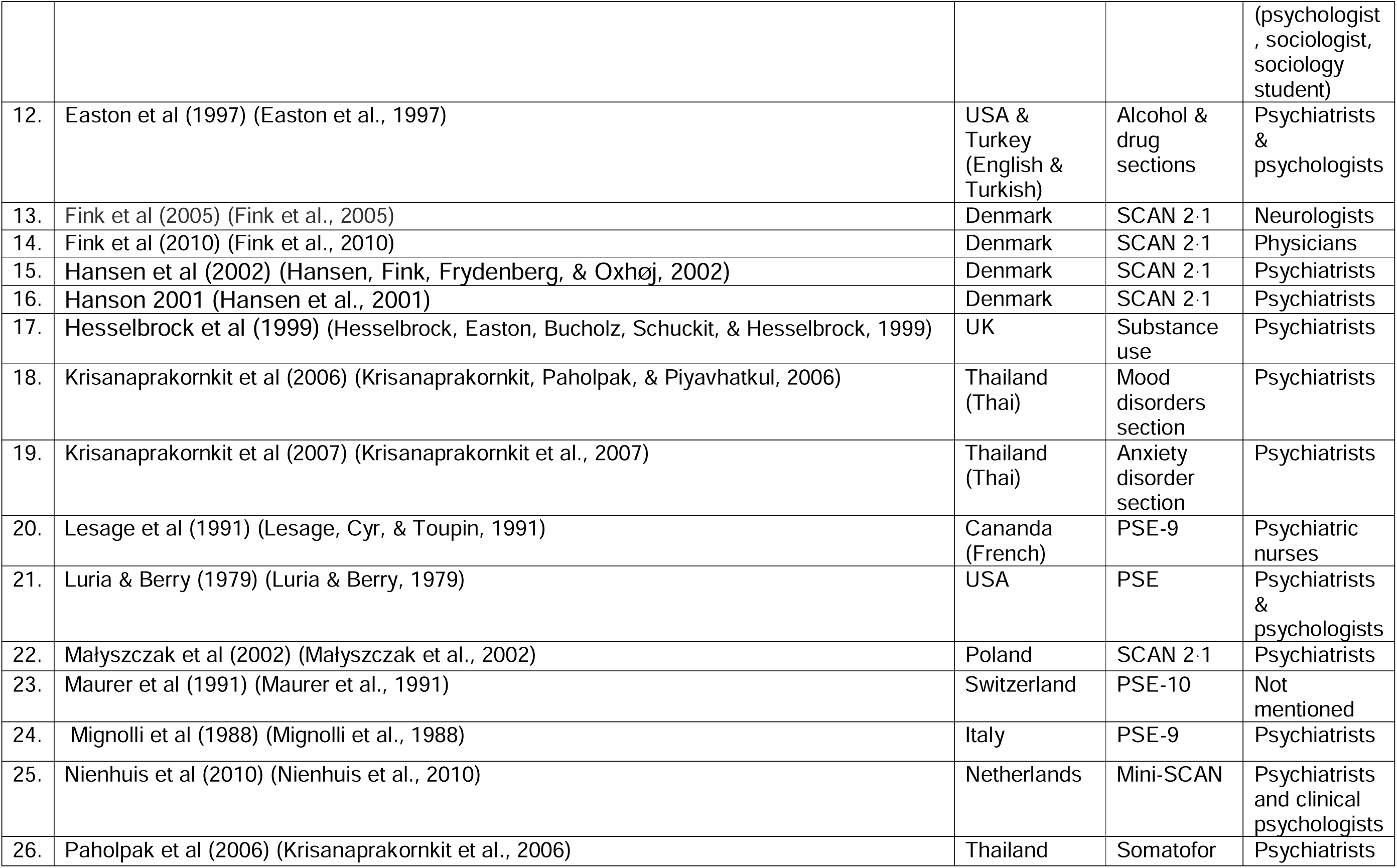
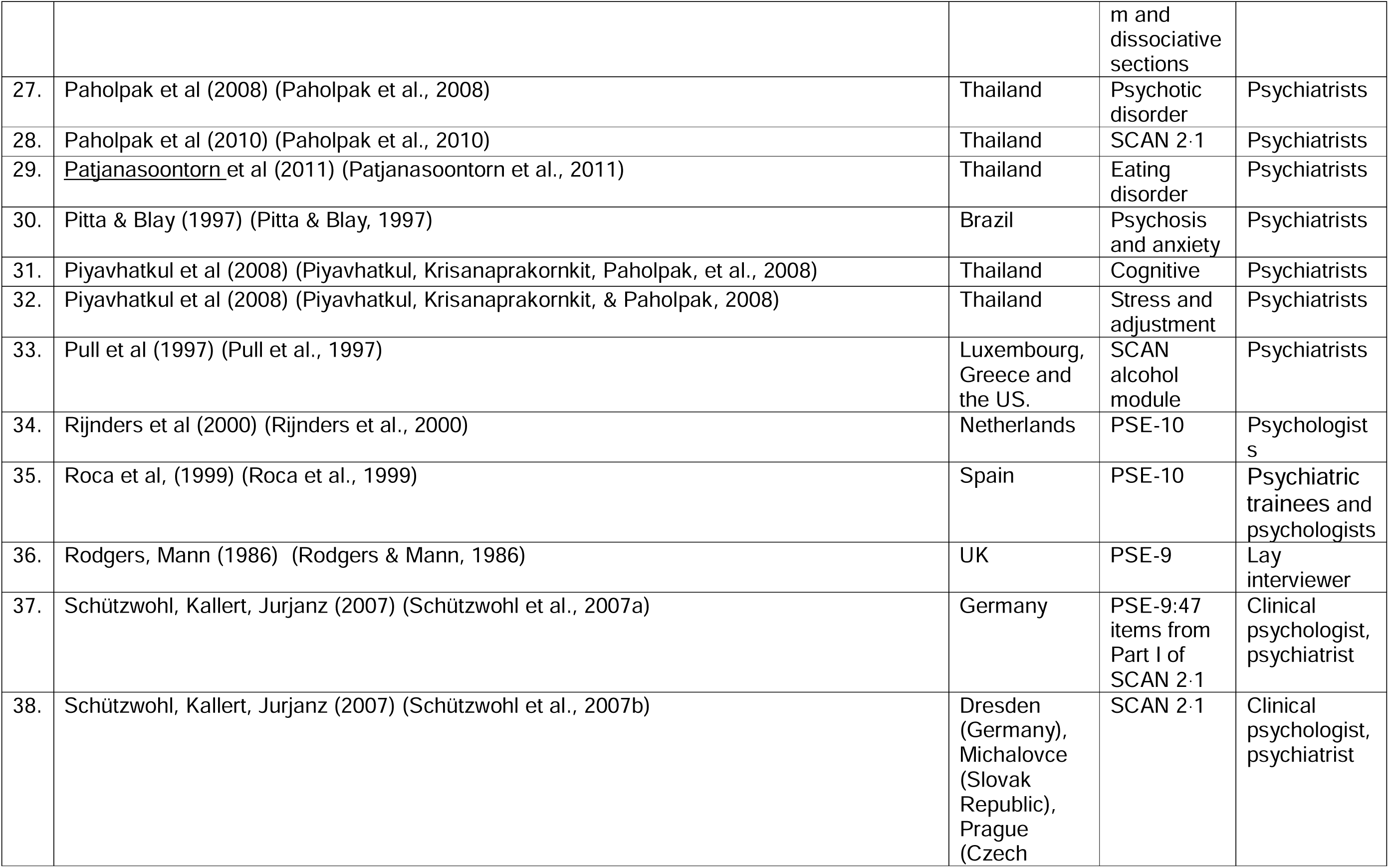
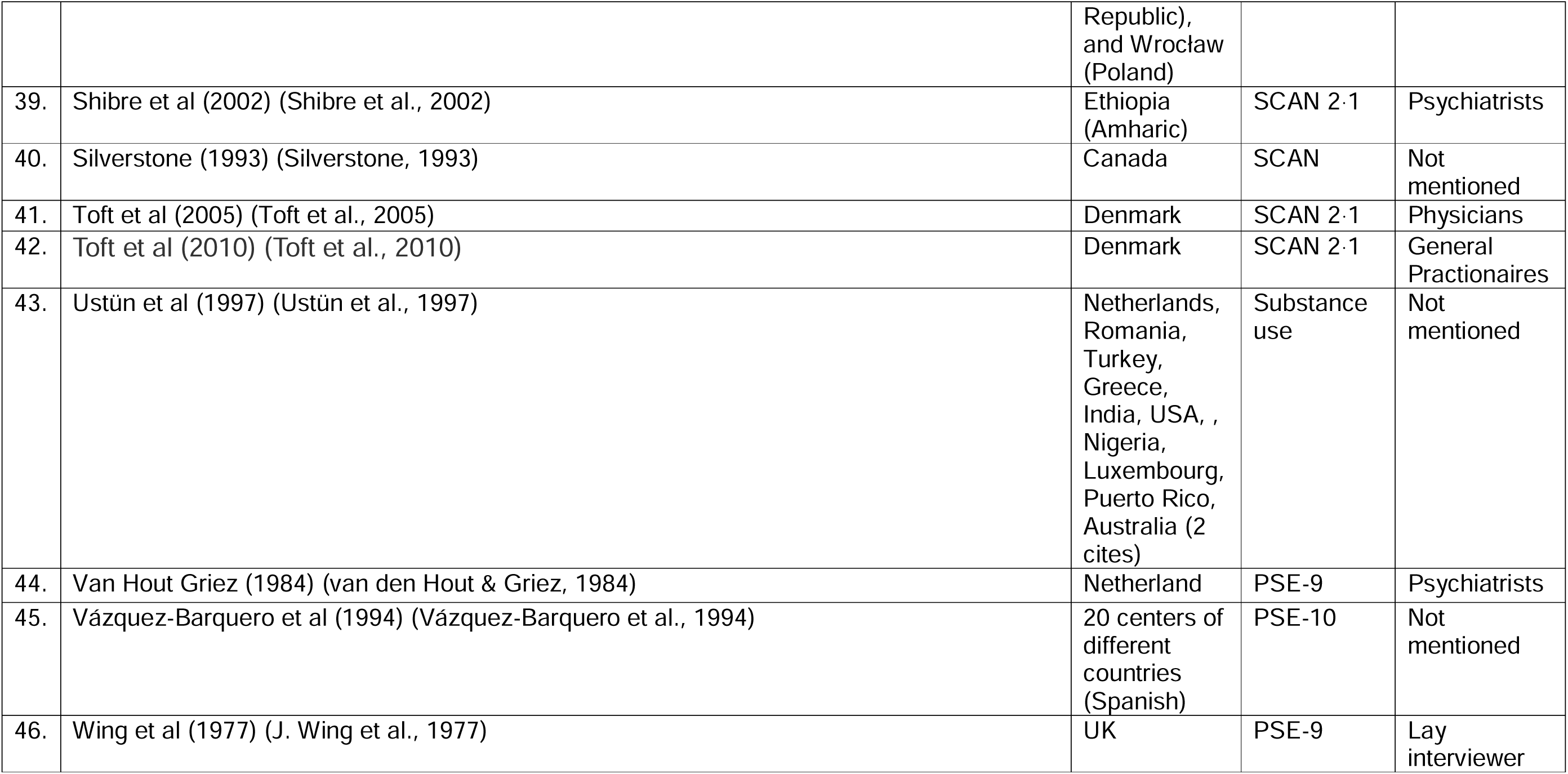
Characteristics of studies examining the psychometric properties of SCAN or PSE.

### 3.3. Bias Assessment

Out of the ten COSMIN risk of bias checklist boxes, we did not rate the first two boxes: development and content validity, as none of the studies we included evaluated these domains for PSE or SCAN.

As most of the studies assessed only inter- and intra-rater reliability, we did not rate the studies for most of the boxes. We completed the checklist for studies that included information on Cronbach’s alpha, test-retest reliability, concurrent validity, and structural validity. The most common issues we identified were low sample sizes, insufficient characterization of the sample, lack of reporting on the time interval between tests, and inadequate description of the comparator instrument used (Supplementary file 1).

### 3.4. Reliability of SCAN or PSE

Our review showed that different versions of the SCAN or PSE were assessed for their reliability, including internal consistency, inter-rater reliability, intra-rater reliability, and test-retest reliability.

In the studies that reported on the internal consistency of SCAN, Cronbach’s alpha values ranged from 0·55 to 0·98 (Schützwohl et al., 2007a; Schützwohl, Kallert, & Jurjanz, 2007b). This indicated good to excellent internal consistency, reflecting the coherence of different domains within SCAN and PSE.

There were also studies (n=8) that examined the intra-rater reliability (instrument administered by the same rater at a prespecified interval, for example, two weeks) of SCAN. These studies reported acceptable agreement across various items or sections of SCAN.

The mean intra-rater kappa value ranged from substantial to almost perfect, although it was slightly lower (but still good) for some sections of SCAN. For somatoform and dissociative symptoms it was at 0.89, anxiety at 0.84, mood at 0.86, eating disorders at 0.76, use of alcohol at 0.82, stress and adjustment disorders at 0.94, psychosis at 0.76, cognitive impairment at 0.78, and observed behaviour, affect, and speech at 0.51. For stress and adjustment disorders and cognitive impairment, the values were between 0.65 and 0.86 (Paholpak, Arunpongpaisal, Krisanaprakornkit, & Khiewyoo, 2008; Paholpak, Arunpongpaisal, Krisanaprakornkit, Piyavhatkul, & Khiewyoo, 2006; Paholpak et al., 2010; Patjanasoontorn, Paholpak, & Krisanaprakornkit, 2011). The intra-class correlation coefficient (ICC) of the SCAN sections ranged from 0·84 to 0·86 (Luria & Berry, 1979).

The other reliability indicator was inter-rater reliability. Overall, there was acceptable agreement in most studies, although not across all settings and mental health conditions. One study indicated a significant level of difference in the parameters across the different mental health conditions: kappa for inter-rater reliability was 0.14 for neurotic disorders and 0.65 for psychotic disorders (Adamowski, Kiejna, & Hadryś, 2006).

Section level inter-rater agreement (kappa value) was also reported: somatoform or dissociative symptoms from 0.77 to 0.88 (Fink, Ørnbøl, & Christensen, 2010; Fink, Steen Hansen, & Søndergaard, 2005), anxiety at 0.79, mood at 0.80, eating disorders at 0.73, use of alcohol at 0.66, stress and adjustment disorders at 0·90, psychosis at 0.68, cognitive impairment at 0.72, and observed behaviour, affect, and speech at 0.45 (Patjanasoontorn et al., 2011). For the substance use disorders section of SCAN, the kappa level of agreement was 0.69 for alcohol, 0.49 for opioids, 0.61 for cocaine and 0.5 for cannabis (Compton, Cottler, Dorsey, Spitznagel, & Mager, 1996).

A high level of inter-rater agreement was observed for the classification of schizophrenia and other psychotic disorders, while a lower level of agreement was found for other mental health conditions. For example, a Kappa value of 0.36 was found for depression, compared to 0.62 for schizophrenia (Małyszczak, Rymaszewska, Hadryś, Adamowski, & Kiejna, 2002). One study reported an overall agreement of 0.81 from live interviews and 0.96 for tape-recorded interviews (J. Wing et al., 1977).

Six studies assessed the test-retest reliability of SCAN or PSE. One study indicated good consistency with a correlation coefficient of 0.67 (J. Wing et al., 1977). The kappa values ranged from 0.7 to 0.9 (Ustün et al., 1997), indicating substantial agreement. Another study reported a kappa level from 0.24 to 0.64 (Rijnders et al., 2000). Test-retest also worked better for psychotic symptoms (kappa =0.82) than non-psychotic symptoms (kappa= 0.16) (J. E. Cooper, Copeland, Brown, Harris, & Gourlay, 1977). There was also a 0.90 kappa for many sections of SCAN (Nienhuis, van de Willige, Rijnders, de Jonge, & Wiersma, 2010).

Regarding substance use disorders, there was acceptable consistency of the SCAN across time for alcohol (0.77-0.85), opiates (0.93-0.98), cannabis (0.63-0.78), and cocaine (0.69-0.74) (Easton et al., 1997). One study indicated a Total Disagreement Index (TDI: total number of disagreements between raters/mean number of positive ratings), and the results were low, indicating the TDI mean is 0.09 (0-0.31) (Mignolli, Faccincani, Burti, Gavioli, & Micciolo, 1988).

#### Validity of SCAN or PSE

Twenty-nine studies reported the concurrent, structural, technical, semantic or language validity of SCAN and PSE.

#### Concurrent validity of SCAN or PSE

Correlation between SCAN and clinical diagnoses using ICD-10 and DSM-IV has been found to vary, ranging from low to strong correlations (Fink et al., 1995; Mignolli et al., 1988; Rijnders et al., 2000; Shibre et al., 2002; Ustün et al., 1997). It had strong or perfect agreement with a clinical diagnosis made by a nurse (Rodgers & Mann, 1986) or a trainee psychiatrist, especially for schizophrenia, but low agreement for depression (Alem, Kebede, Shibre, Negash, & Deyassa, 2004; Małyszczak et al., 2002). Another study showed that SCAN demonstrated 100% agreement for schizophrenia, 95.3% for bipolar disorder, and 93% for depression when compared to clinical diagnoses (Alem et al., 2004).

One study reported a sensitivity of 0.38 and specificity of 0.91 for physicians at admission and a sensitivity of 0.48 and specificity of 0.87 for nurses at admission. At discharge, the sensitivity was 0.40 and the specificity was 0.89 for physicians, while for nurses, the sensitivity was 0.56 and the specificity was 0.81 (Hansen et al., 2001).

The sensitivity and specificity for the psychosis section of SCAN were notably higher at 0.69 and 0.94, respectively. However, specificity was lower for other modules, such as affective disorders (0.41) and neurotic disorders (0.5) (Adamowski et al., 2006). The sensitivity against ICD-10 diagnoses from various studies ranged from 0.69 to 0.95, while specificity ranged from 0.41 to 0.94 (Adamowski et al., 2006; Brugha, Bebbington, et al., 1999)

#### Construct validity of SCAN or PSE

Convergent validity between SCAN and mini-SCAN demonstrated substantial agreement (Nienhuis et al., 2010; Pull et al., 1997). Structural validity, or the extent to which SCAN and PSE scores adequately reflect the dimensionality of the constructs they aim to measure, has also been examined using exploratory factor analysis.

For the PSE, the findings indicated a unidimensional structure. Many of the PSE items had high loading on the dominant factor (Schützwohl et al., 2007b), but the shared variance was small (Rodgers & Mann, 1986). The first extracted factor accounted for 20.3% of the total variance, but the values of the second, third, and fourth factors accounted for only 5.5%, 4.5%, and 3.7%, respectively (Rodgers & Mann, 1986). The exploratory factor analysis further indicated that SCAN effectively captures major psychiatric dimensions. In section 10, the mood disorder section factor loading ranged from 0.88 to 0.97, the depression section with a factor loading ranged from 0.89 to 0.92, and the thinking section ranged from 0.24 to 0.88 (Schützwohl et al., 2007b). There was also substantial agreement between the different sections of SCAN and its total scores (Pull et al., 1997).

#### Other properties

The SCAN and PSE have also demonstrated flexibility in administration, including duration of administration and practicality. The studies indicated that the duration of administration fluctuates based on the complexity of the condition and the specific section (Mignolli et al., 1988) being administered. The mini-SCAN interview takes about 25 minutes less than the full SCAN (Nienhuis et al., 2010).

The alcohol module alone took about 40 minutes to administer, the stress-related disorders module took about 26 minutes (Piyavhatkul, Krisanaprakornkit, & Paholpak, 2008), and 60 minutes for people with cognitive impairment (Piyavhatkul, Krisanaprakornkit, Paholpak, & Khiewyou, 2008). On average, it took 57 minutes for somatoform and dissociative disorders (Paholpak et al., 2006).

In all the studies and all sections of SCAN, administering SCAN to people with impairment took longer compared to the healthy population. This was particularly the case among people with severe mental health conditions. Administering SCAN to people with schizophrenia took 140 ± 36.0 minutes, while it only took 82 ± 25.9 minutes (range 48-124 minutes for the healthy population (Paholpak et al., 2008).

Two studies have indicated the acceptability of SCAN (Brugha, Bebbington, et al., 1999; Nienhuis et al., 2010). Nienhuis et al. (2010) reported that 77% and 79% of the participants rated the SCAN and mini-SCAN interviews, respectively, as pleasant or very pleasant (Nienhuis et al., 2010). There were minimal reported difficulties in administering SCAN, especially when experience builds.

Studies that reported content, semantics, languages, and content validity indicated that SCAN could be effectively translated into different languages. There were no significant concerns reported (Table 2).

**Table 2:**
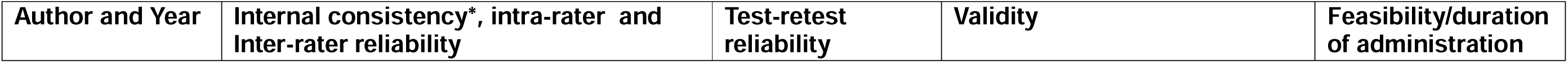
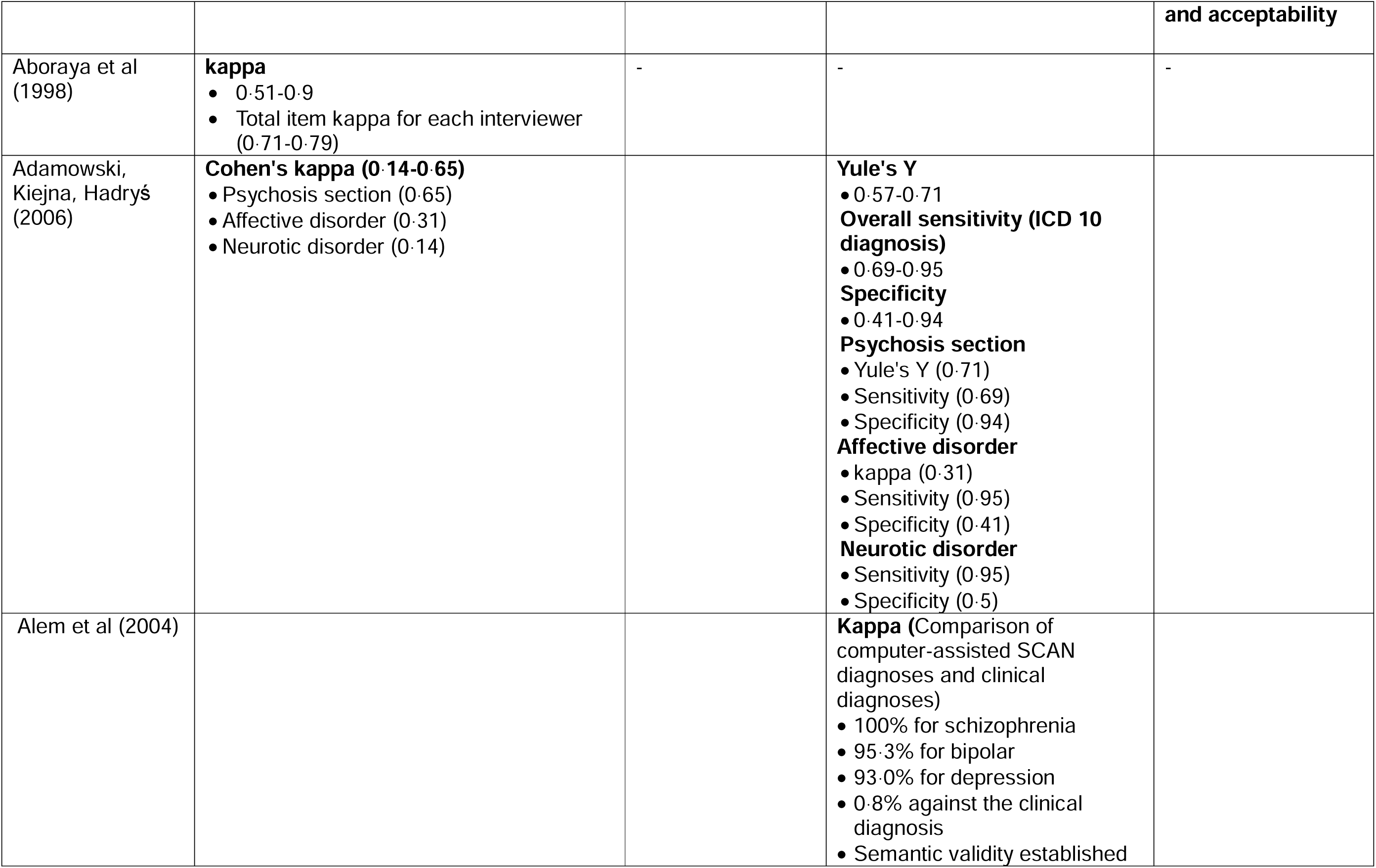
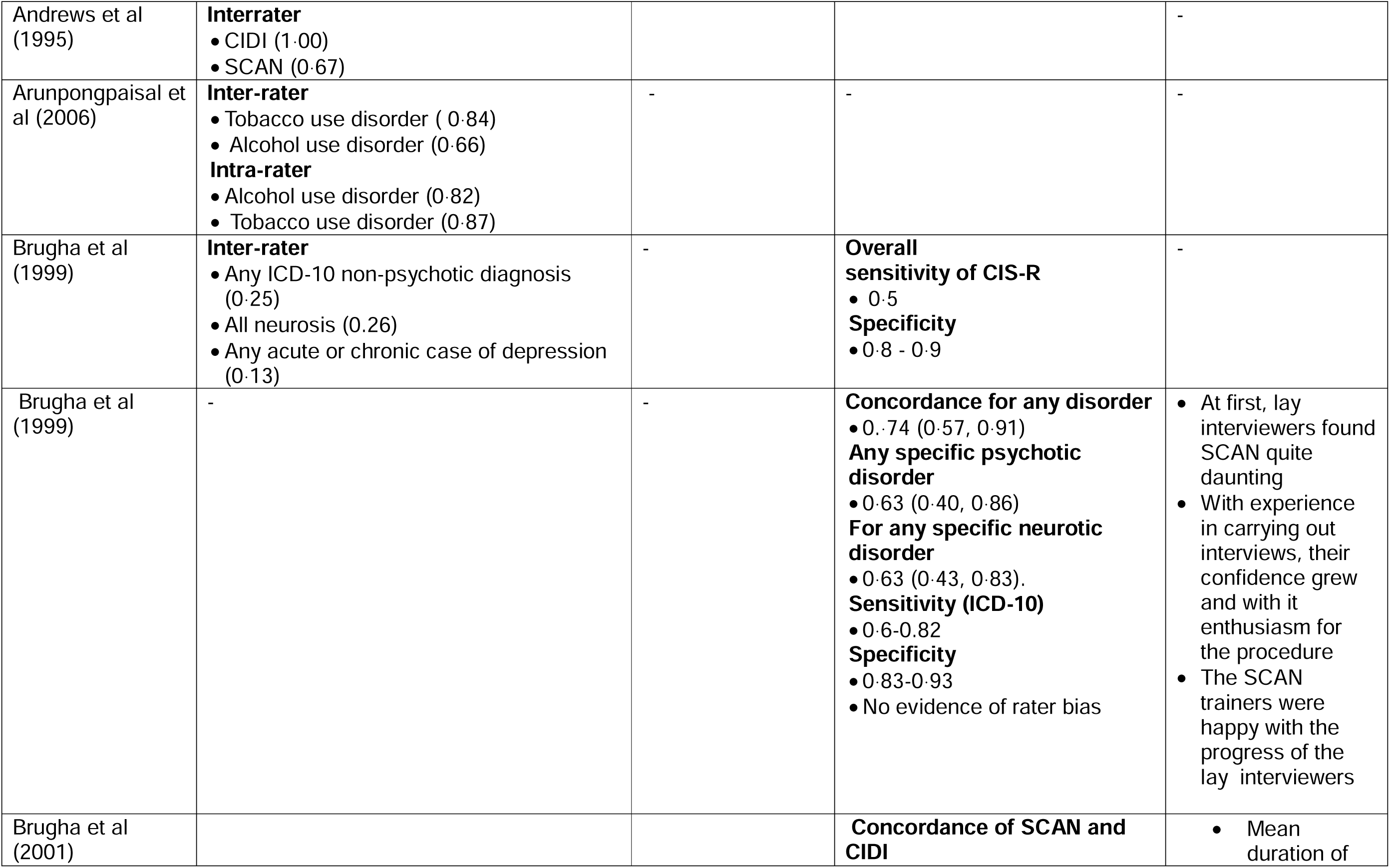
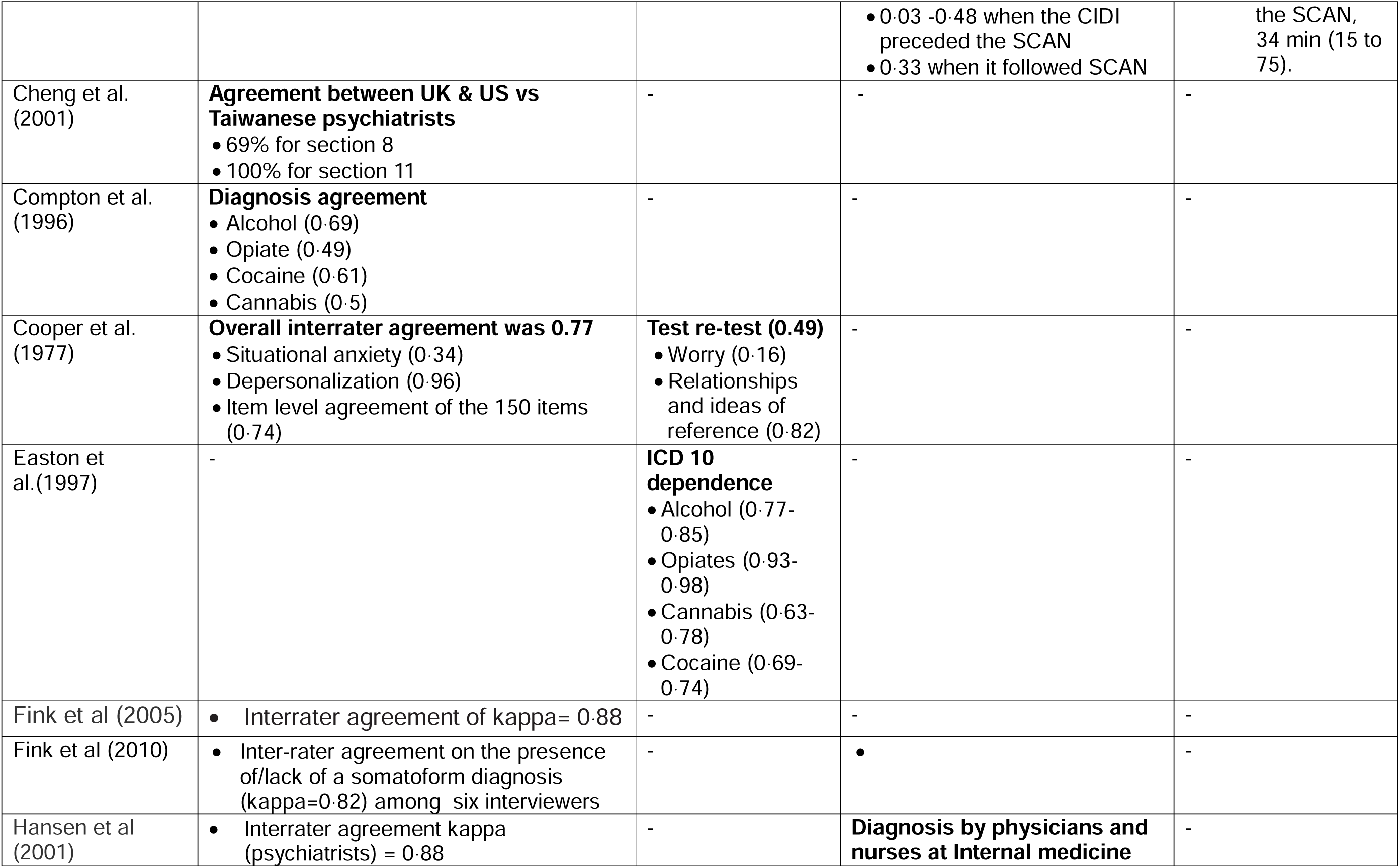
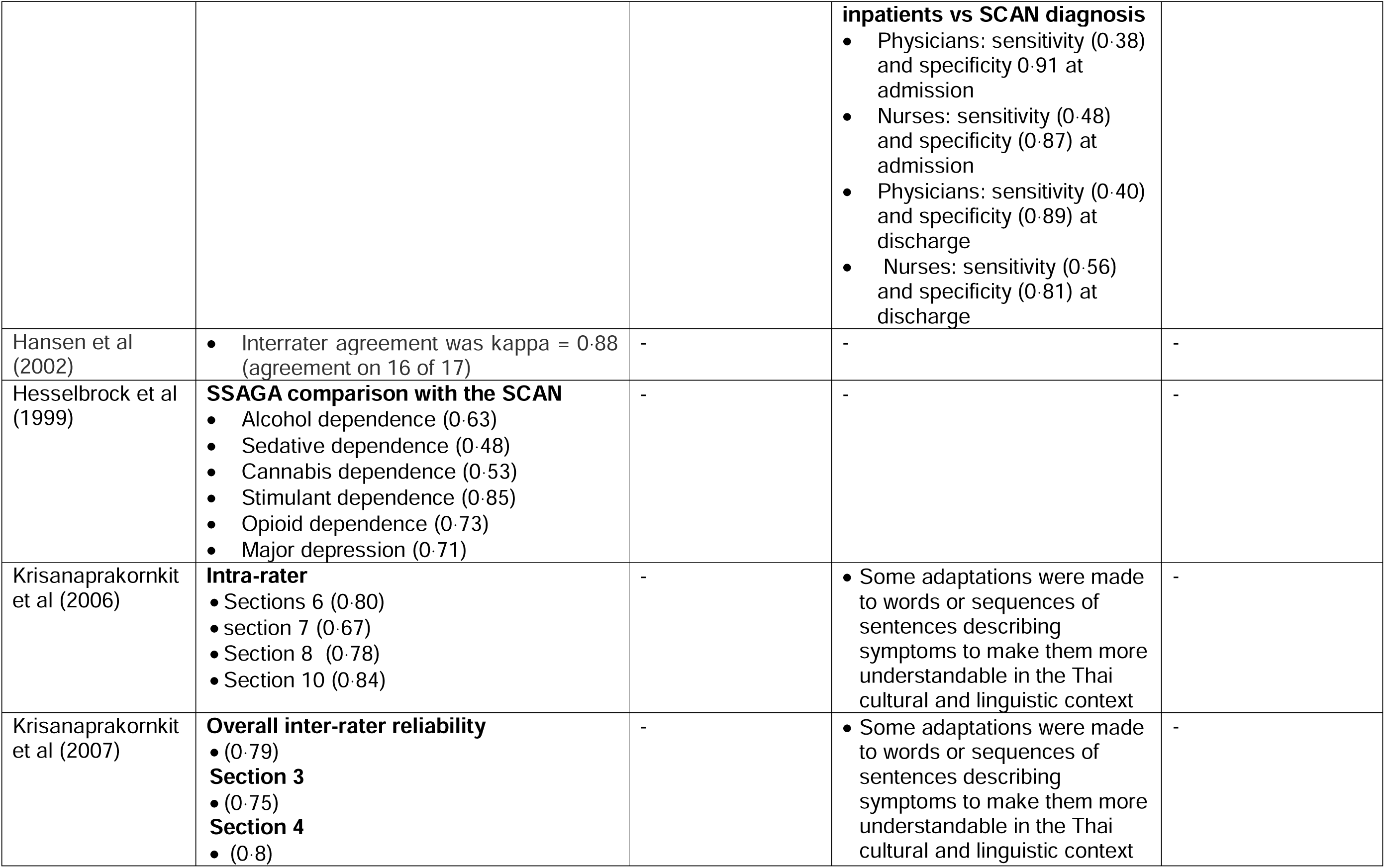
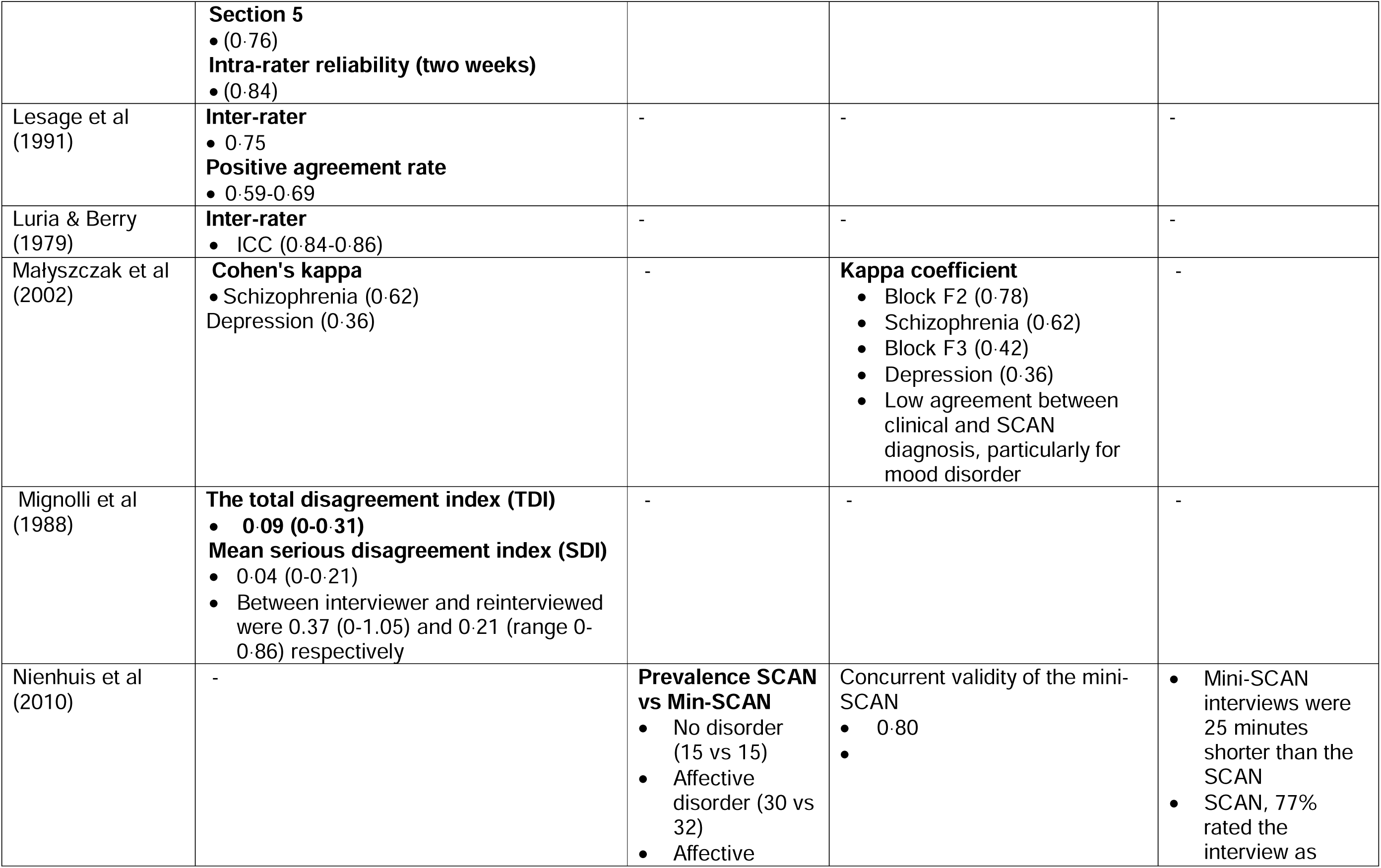
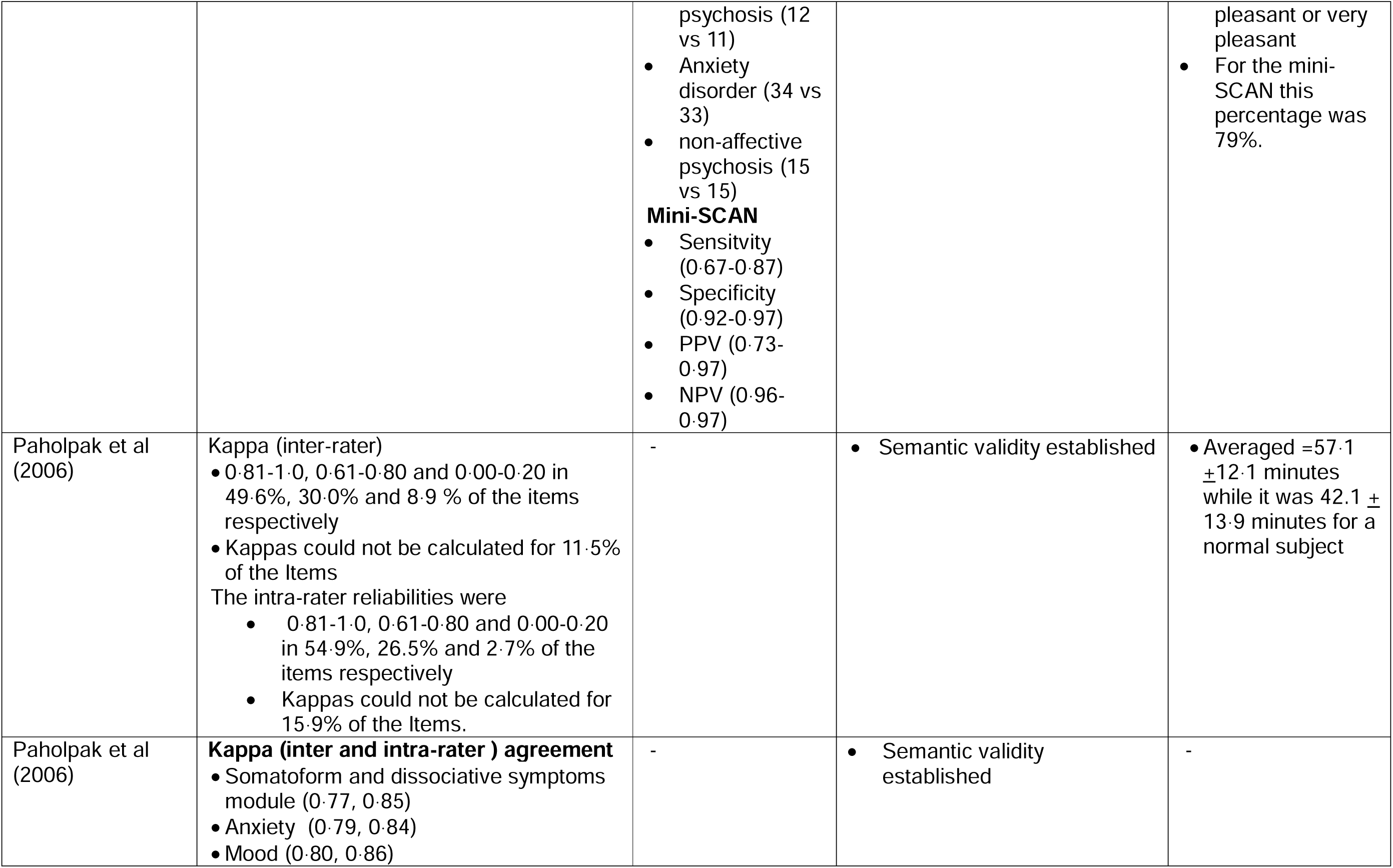
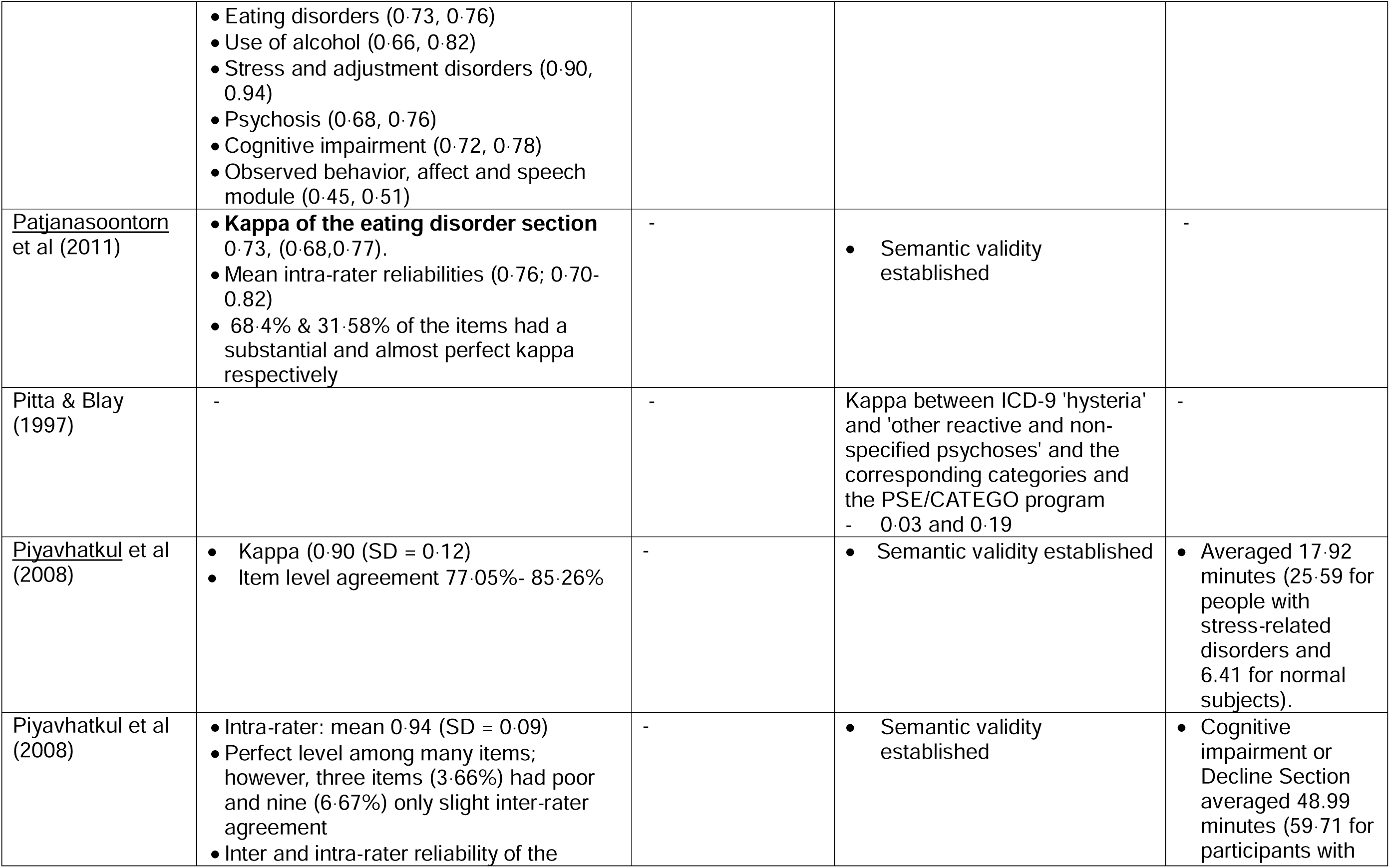
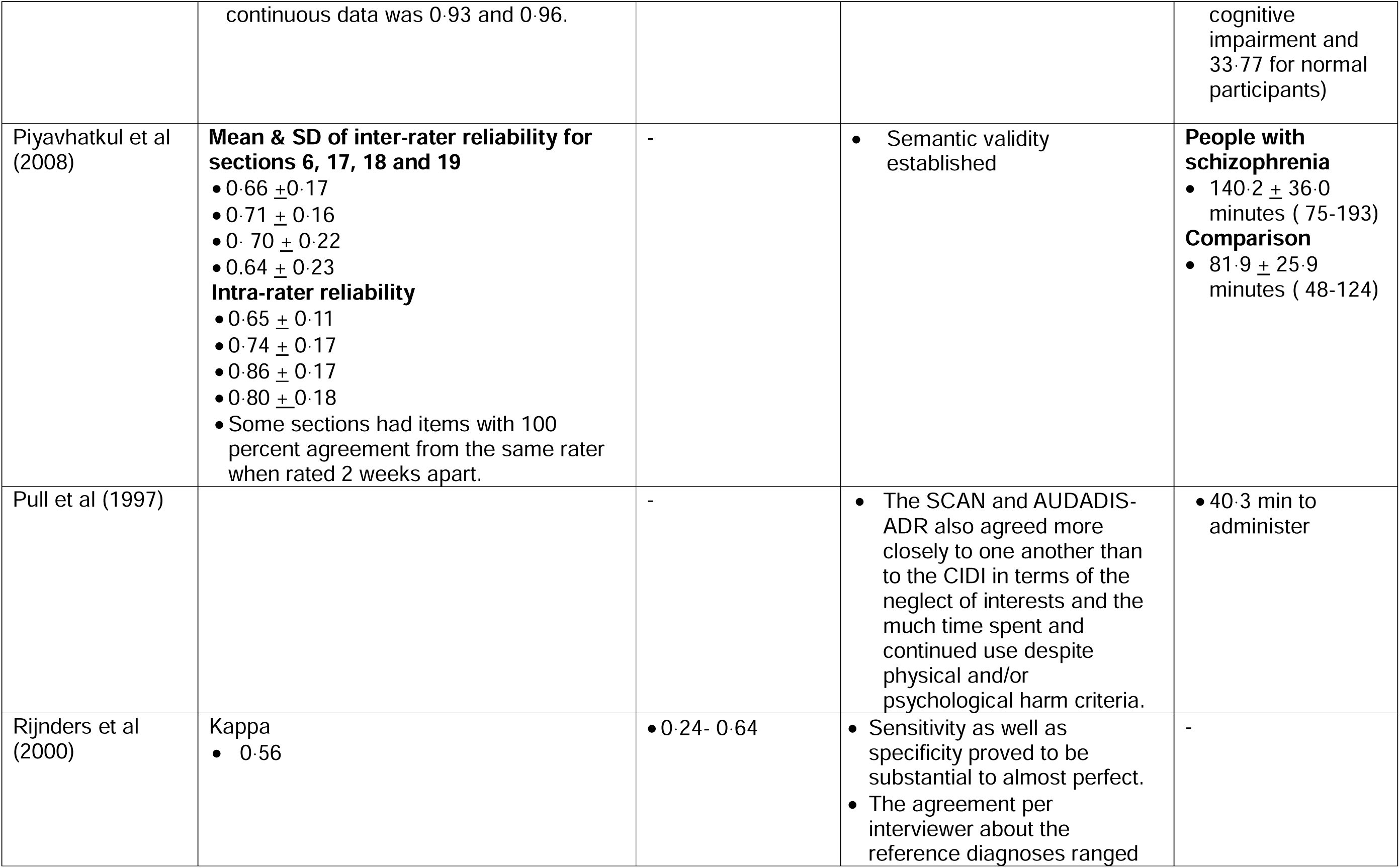
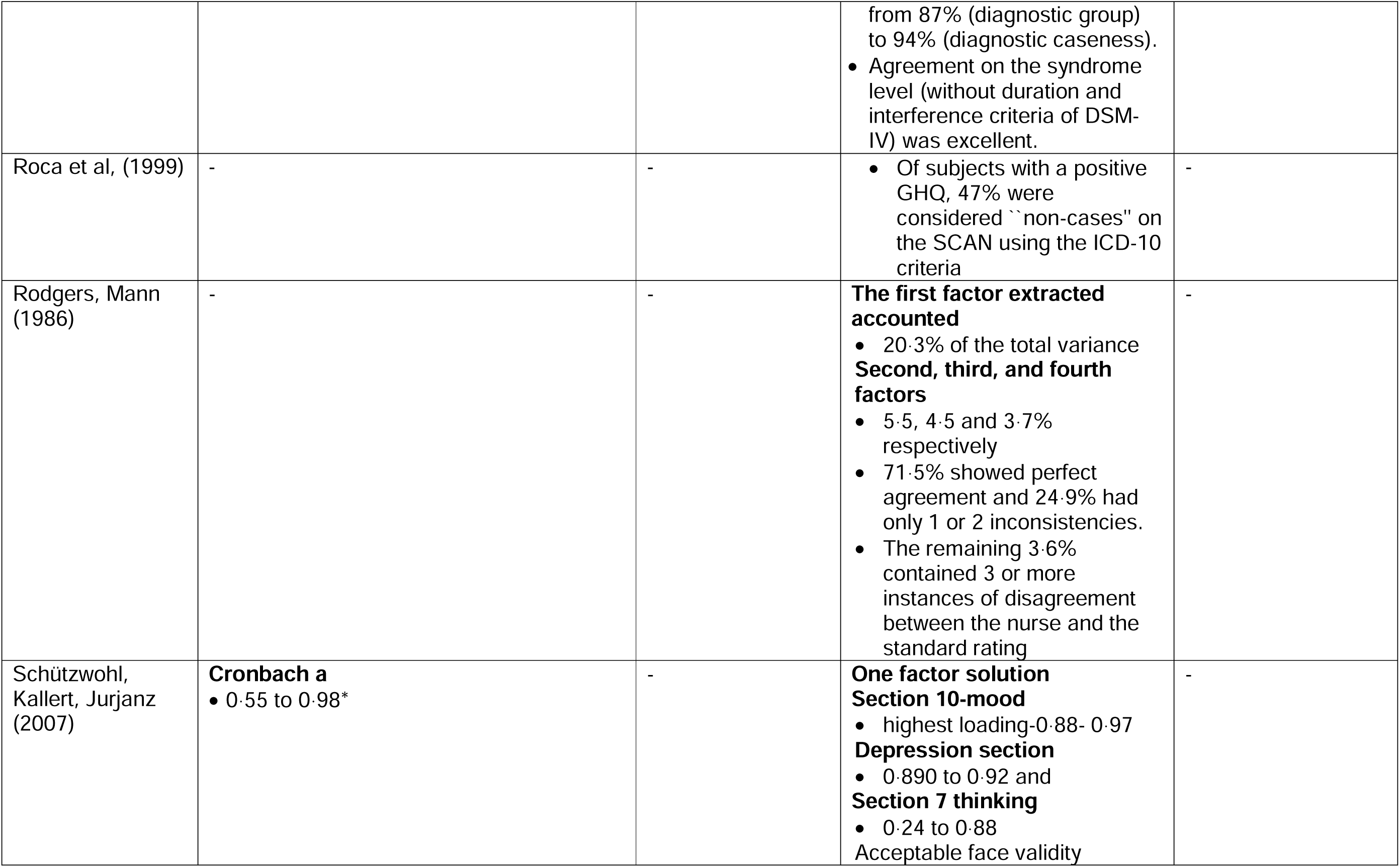
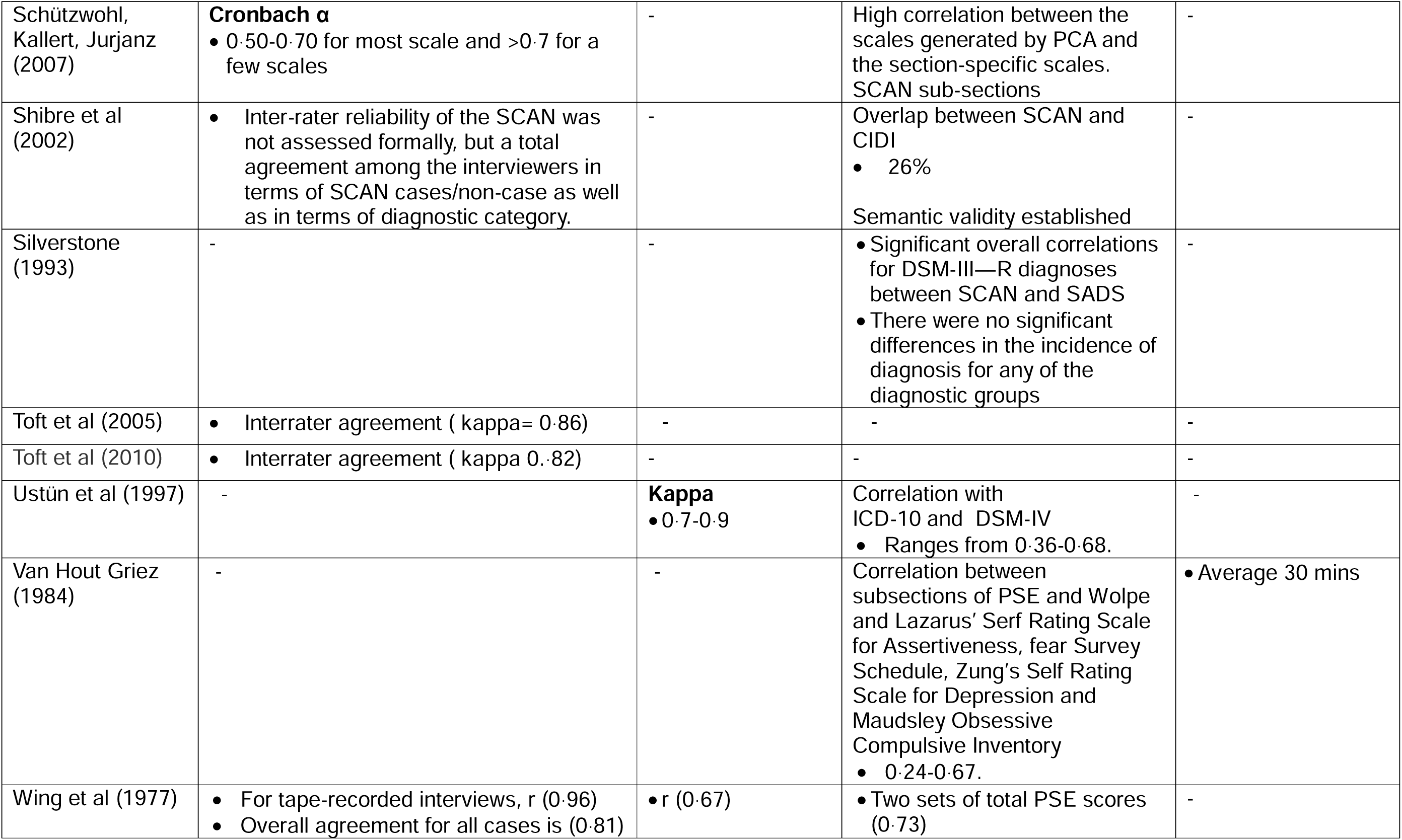
Summary of psychometric properties of SCAN or PSE.

### 3.5. SCAN or PSE as a gold standard

Forty-seven articles used SCAN or PSE versions to evaluate the psychometric properties of other diagnostic or screening tools. The publication year ranged from 1989 to 2023. Most of the studies, except for five (Ethiopia (n=3), Nigeria, and South Africa), were conducted in high-income countries.

A total of 31 tools were validated using SCAN or PSE as a gold standard. Of the 31, 18 were used to assess depression, while the other measures were used to assess symptoms of psychosis, anxiety, somatoform, and substance use problems. Some of the scales or assessment instruments (n=4), such as the CIDI, were used to identify more than one psychiatric problem. These 31 measures were validated in more than 15 different languages (Supplementary file 2).

## 4. Discussion

We aimed to systematically review the psychometric properties of the various revisions of the SCAN. We found 46 original psychometric papers on SCAN and PSE and another 47 articles that used SCAN as a gold standard to evaluate the psychometric properties of other mental health measures. Most of the studies used multiple sections of SCAN, while some used one or two sections out of the 27 sections.

The internal consistency of the items in SCAN and PSE ranged from good to excellent. Additionally, both modules demonstrated acceptable intra-rater, inter-rater, and test-retest reliability. However, some studies reported percentages and mean (Cheng et al., 2001; Compton et al., 1996; Piyavhatkul, Krisanaprakornkit, Paholpak, et al., 2008) to calculate various types of reliabilities. This might introduce methodological biases and require cautious interpretation.

There was also evidence supporting the concurrent, semantic, and content validity of both SCAN and PSE, though there was no evidence for predictive validity. The good to excellent internal consistency of SCAN and PSE suggests that the items effectively measure the underlying construct of psychopathology. But, internal consistency should have been calculated for each section of the SCAN or PSE, not the total score (Lidwine B Mokkink et al., 2012). Another issue to consider is that the high internal consistency of the SCAN and PSE might be attributed to the large number of items in these scales, rather than their actual reliability (Meyer et al., 2001). High internal consistency is not an ideal indicator for tools with many items (Lidwine B Mokkink et al., 2012).

We also found acceptable agreement across various items or sections of SCAN and PSE. However, methodological concerns arise from the use of percentage agreement and Cohen’s Kappa when the studies calculated test-retest, intra-rater, and inter-rater reliability. Unlike Kappa, percentage agreement does not account for the possibility that agreement may occur by chance.

Additionally, high agreement among raters on specific sections, particularly those related to schizophrenia or psychotic disorders, could positively influence the overall percentage of agreement when considering the entire SCAN and PSE. Only a few studies calculated the ICC, which is a rigorous methodology that considers total variance, including between participants, between raters, and residual error. However, these studies did not report measurement errors (Lidwine B Mokkink et al., 2012).

Although the evidence is not robust, both SCAN and PSE appear more reliable for psychotic disorders, which present with more severe, clear, and observable symptoms compared to neurotic or common mental disorders. Of interest, the strengths and weaknesses of the SCAN and instruments such as CIDI are inverse, as CIDI does not perform well for psychotic disorders (L. Cooper, Peters, & Andrews, 1998). This suggests that improving the sections on common mental disorders in the SCAN could be a focus for future research.

As the PSE and SCAN have been translated into many languages, considerable experience has been gained. Translating concepts, not just words, and the flexibility of SCAN, allowing administrators to explain and probe, suggests its strong semantic validity (J. K. Wing et al., 1990). Studies also showed that they were able to establish that the back-translation of SCAN and PSE into the original English is appropriate.

Because SCAN is typically considered a gold standard, criterion validity has been difficult to study. Few studies have reported acceptable sensitivity and specificity of SCAN in comparison to diagnosis by clinicians, particularly for psychotic disorders as opposed to affective disorders (Adamowski et al., 2006).

The positive and negative predictive power of SCAN remains uncertain, as it has not been extensively studied. Correlations between SCAN and diagnosis using ICD-10 and DSM-IV were found to vary, ranging from low to strong correlation (Fink et al., 1995; Rijnders et al., 2000; Shibre et al., 2002; Ustün et al., 1997).

The semi-structured nature of SCAN allows flexibility for the test administrator to rephrase and probe, and the common practice of its use by highly trained mental health professionals could potentially contribute to better validity compared to other structured diagnostic tools, such as the CIDI (Brugha, Jenkins, Taub, Meltzer, & Bebbington, 2001).

There is a need for strong studies that report the construct validity of SCAN using factor analysis. There is only such a study which reported one dominant factor and high factor loading of the items (Schützwohl et al., 2007b). This finding is like studies on CIDI and other diagnostic tools, which tend to be unidimensional per section (Gelaye et al., 2013). Further research on the cultural invariance of SCAN is warranted, as this would facilitate cross-cultural comparisons of psychiatric disorders.

SCAN is accepted and used globally (Gureje et al., 2020; Hanlon et al., 2016; Hanlon et al., 2024; Roberts et al., 2020). Previous studies have also noted that SCAN can be effectively utilized by trained and experienced lay interviewers (Brugha, Bebbington, et al., 1999). The computer algorithm for interpretation and its dimensional measurement capabilities are additional strengths of SCAN (WHO, 1994).

## Conclusion

Considering the methodological weakness of the studies that reported the psychometric properties of SCAN when evaluated against recently recommended standards, there is a clear need for further studies. Additionally, we couldn’t find recent studies that evaluate the psychometric properties of SCAN.

Despite these limitations, SCAN continues to be a promising tool in terms of some of its psychometric properties and practical feasibility for diagnosing psychiatric disorders. Its established semantic, content, along with some accepted reliability indicators, underscore its potential as a valuable diagnostic instrument.

The next version of SCAN must consider its potential limitations, particularly in the non-psychotic sections. Additionally, it is important to revise the current version considering the ICD-11 and DSM-5 diagnostic systems, as the current version is based on the previous versions.

## Contributions

MO, AF, AA, and WF conceived the study. WF, AM, AF, and AA contributed to the design of the review and the protocol development. WF and AM search the literature, data collection, and quality appraisal. All authors participated in contacting researchers who used SCAN and in retrieving some of the old publications. WF and AM drafted the manuscript. All authors revised the manuscript and approved the final manuscript.

## Availability of Data and Materials

The authors confirm that the data supporting the findings of this study are available in the article and its supplementary materials.

## Financial declarations

WF and AM received support from WHO. WF and CH receive support from the Wellcome Trust through grants 222154/Z20/Z. CH receives support from NIHR through the NIHR Global Health Research Group on Homelessness and Mental Health in Africa (NIHR134325), using UK aid from the UK Government. CH also receives support from WT grant 223615/Z/21/Z. AM receives support from DELTAS Africa Initiative through the African Mental Health Research Initiative (AMARI) II project. The DELTAS Africa Initiative is a programme of the Science for Africa Foundation being implemented with support from Wellcome and the UK Foreign Commonwealth and Development Office (FCDO). The views expressed are those of the authors and not necessarily those of the NIHR, the Department of Health and Social Care or WHO.

For the purpose of open access, the authors have applied a Creative Commons Attribution (CC BY) licence to any Author Accepted Author Manuscript version arising from this submission.

